# Assessment of machine learning algorithms to predict medical specialty choice

**DOI:** 10.1101/2025.03.06.25323485

**Authors:** David Vicente Alvarez, Milena Abbiati, Alban Bornet, Georges Savoldelli, Nadia Bajwa, Douglas Teodoro

## Abstract

Equitable distribution of physicians across specialties is a significant public health challenge. While previous studies primarily relied on classic statistics models to estimate factors affecting medical students’ career choices, this study explores the use of machine learning techniques to predict decisions early in their studies. We evaluated various supervised models, including support vector machines, artificial neural networks, extreme gradient boosting (XGBoost), and CatBoost using data from 399 medical students from medical faculties in Switzerland and France. Ensemble methods outperformed simpler models, with CatBoost achieving a macro AUROC of 76%. Post-hoc interpretability methods revealed key factors influencing predictions, such as motivation to become a surgeon and psychological traits like extraversion. These findings show that machine learning could be used for predicting medical career paths and inform better workforce planning.

## 1. Introduction

The Global Health Workforce Alliance identifies the unequal distribution of medical personnel as one of the foremost public health challenges of the 21st century [1]. Ensuring an equitable allocation of physicians across various specialties and geographic regions remains a significant challenge for developed nations, many of which rely heavily on internationally trained physicians to meet their healthcare needs [2,3]. Addressing this issue necessitates a coordinated effort to ensure that enough qualified and appropriately specialized physicians are strategically placed to serve regions that are currently underserved.

Medical student career choice intentions and motives have a direct effect on physician workforce planning. Individual factors, such as gender, personality, and motivation, have been identified as significantly impacting the dynamics of the career choice process [4]. Existing studies have focused mostly on identifying factors associated with career choice rather than predicting student career intentions [5–7]; with methods based on classic statistics, such as logistic regression. To the best of our knowledge, only one study applied machine learning methods to predict medical student’s career choices [8]. However, it focused only on artificial neural networks (ANN). However, ANN tends not to be the best modeling choice for tabular data, being often outperformed by gradient-boosting methods, such as extreme gradient boosting (XGBoost) [9]. To address this gap, in this study, we aimed to assess several machine learning algorithms for predicting medical students’ career choices. Another contribution of this work is that we investigated post-hoc interpretability methods to identify the factors influencing students’ specialty selection, which could provide hypotheses for factors influencing the career decision process.

## 2. Methods

### 2.1. Career choice dataset

The career choice dataset contained socio-educational, psychological [10,11], and motivational features for 686 (male=244; female=441) medical students enrolled at the Universities of Geneva (n=285; 41.6%, male=110; female=175), Lausanne (n=141; 20.5%, male=43; female=98), and Strasbourg (n=260; 37.8%, male=91; female=168). Students completed the questionnaire twice: once at the start (year 4) and once at the conclusion of their shared studies (year 6). The outcome labels were defined as the choice of the specialty made by the student in their 6^th^ year. For simplicity, these labels were separated into 2 by domain experts: “technically oriented specialties” and “person-oriented specialties”: Any missing data point was coded as -1. After data cleaning, including the removal of instances with missing labels or duplicates, the dataset comprised 399 (male=130; female=269) instances.

### 2.2. Experiment

We trained several machine learning classifiers [12] - logistic regression, decision tree, k-nearest neighbors, support vector machine (SVM), random forest, multi-layer perceptron (MLP), XGBoost, and CatBoost - using a 5-fold cross-validation scheme. In every fold, we used 60% for training, 20% hyper-parameter fine-tuning, and 20% for testing. For hyper-parameter fine-tuning, we did a grid search and used the parameters that led to the best F1-score over the dataset. We then compared model performances against a majority-class classifier, which naively predicted the majority class from the training dataset. We evaluated all models using the testing dataset. We report the average and standard deviation across all folds for the F1-score, area under the receiver operating characteristic curve (AUROC), and accuracy metrics. We used SHAP (SHapley Additive exPlanations) [13] values to assess the relative importance of different features in predicting medical student career choices.

## 3. Results and Discussions

Table 1 shows the results of our experiments. Among the more sophisticated models, CatBoost and MLP perform comparably, with CatBoost exhibiting the highest AUROC (75.7%) and accuracy (69.2%) while MLP achieved the highest F1-score (67.8%). XGBoost, another gradient-boosting model, performs competitively, with an F1-score of 66.5%, an AUROC of 75.0%, and an accuracy of 68.9%. Despite its simpler computational complexity, the performance of logistic regression is relatively strong, with an F1-score of 64.6% and AUROC of 72.7%, closely followed by the SVM. Random forest has a decent AUROC of 71.4%, however, its overall accuracy and F1-score are lower compared to other models. The kNN and decision tree classifiers are outperformed by most other models. While kNN shows the lowest performance among the non-baseline models, with an F1-score of 52.5% and AUROC of 57.1%, decision tree has moderate success with an F1-score of 56.0% and AUROC of 56.6%. These results indicate that ensemble machine learning algorithms, particularly XGBoost and CatBoost, along with MLPs, outperform simpler models such as decision trees and kNN in predicting the career choices of medical students. The higher performance of ensemble methods is consistent with their well-documented strengths in handling structured tabular data [14].

**Table 1.** Performance evaluation of all networks

| Network | Macro F1-score (std) | Macro AUROC (std) | Accuracy (std) |
| --- | --- | --- | --- |
| Majority vote classifier | 36.7% ($\pm 2.2\%$) | 50.0% ($\pm 0.0\%$) | 58.1% ($\pm 5.4\%$) |
| Logistic regression | 64.6% ($\pm 4.2\%$) | 72.7% ($\pm 2.8\%$) | 65.2% ($\pm 4.5\%$) |
| KNN | 52.5% ($\pm 3.5\%$) | 57.1% ($\pm 6.6\%$) | 56.1% ($\pm 3.8\%$) |
| Decision tree | 56.0% ($\pm 5.2\%$) | 56.6% ($\pm 5.5\%$) | 57.4% ($\pm 4.8\%$) |
| SVM | 63.0% ($\pm 4.3\%$) | 71.2% ($\pm 1.9\%$) | 63.4% ($\pm 4.6\%$) |
| Random forest | 60.9% ($\pm 4.6\%$) | 71.4% ($\pm 4.3\%$) | 63.4% ($\pm 4.4\%$) |
| MLP | **67.8% ($\pm 2.9\%$)** | 72.1% ($\pm 3.6\%$) | 68.9% ($\pm 3.6\%$) |
| XGBoost | 66.5% ($\pm 5.0\%$) | 75.0% ($\pm 5.4\%$) | 68.9% ($\pm 4.6\%$) |
| CatBoost | 67.6% ($\pm 3.9\%$) | **75.7% ($\pm 3.7\%$)** | **69.2% ($\pm 3.9\%$)** |

### 3.1. Explainability

Due to the out-of-the-box implementation of interpretability functionalities via SHAP values and the close performance to the best model, CatBoost, we used the XGBoost model to identify features impacting the decision of the model for career prediction. Figure 1 shows the SHAP values for the XGBoost classifier. The left plot summarizes feature importance, while the right plot highlights the impact of features on predicting a student’s choice of “technically oriented specialties.” The Y-axis ranks features by importance, while the X-axis shows SHAP values: positive values push predictions toward choosing a technically oriented specialty, and negative values push them away, with the color gradient indicating feature values (blue = low, pink/red = high). Features such as motivation for becoming a surgeon (“MOT SURGEON”), motivation for general practice (“MOT GENERAL PRACTIONNER”), and the level of the profession of the father (“PROFESSION FATHER”) have the most substantial influence on the model’s output, with higher SHAP values indicating a greater impact. Moreover, high values of motivation for becoming a surgeon push the model’s prediction towards the “technically oriented specialties” class (positive SHAP values), while high values of motivation for becoming a general practitioner decrease the likelihood (negative SHAP values) of choosing a technically oriented specialty. Psychological features like openness (“NEOO”), extraversion (“NEOE”), and self-determined motivation (AMQTOT) play also a relevant role in predicting career decisions. When a high value for the extraversion score is present, the model tends to predict the “technically oriented specialties” class. On the other hand, a high value for openness or self-determined motivation scores pushes the model to predict toward the “person-oriented specialties” class. The educational level of parents (“LEVEL EDU MOTHER” and “LEVEL EDU FATHER”), practice choice (“PRACTICE CHOICE” *private vs public domain*), and gender (“SEX”) show limited influence on the model’s decision, which is interestingly divergent from current studies [4].

**Figure 1.**
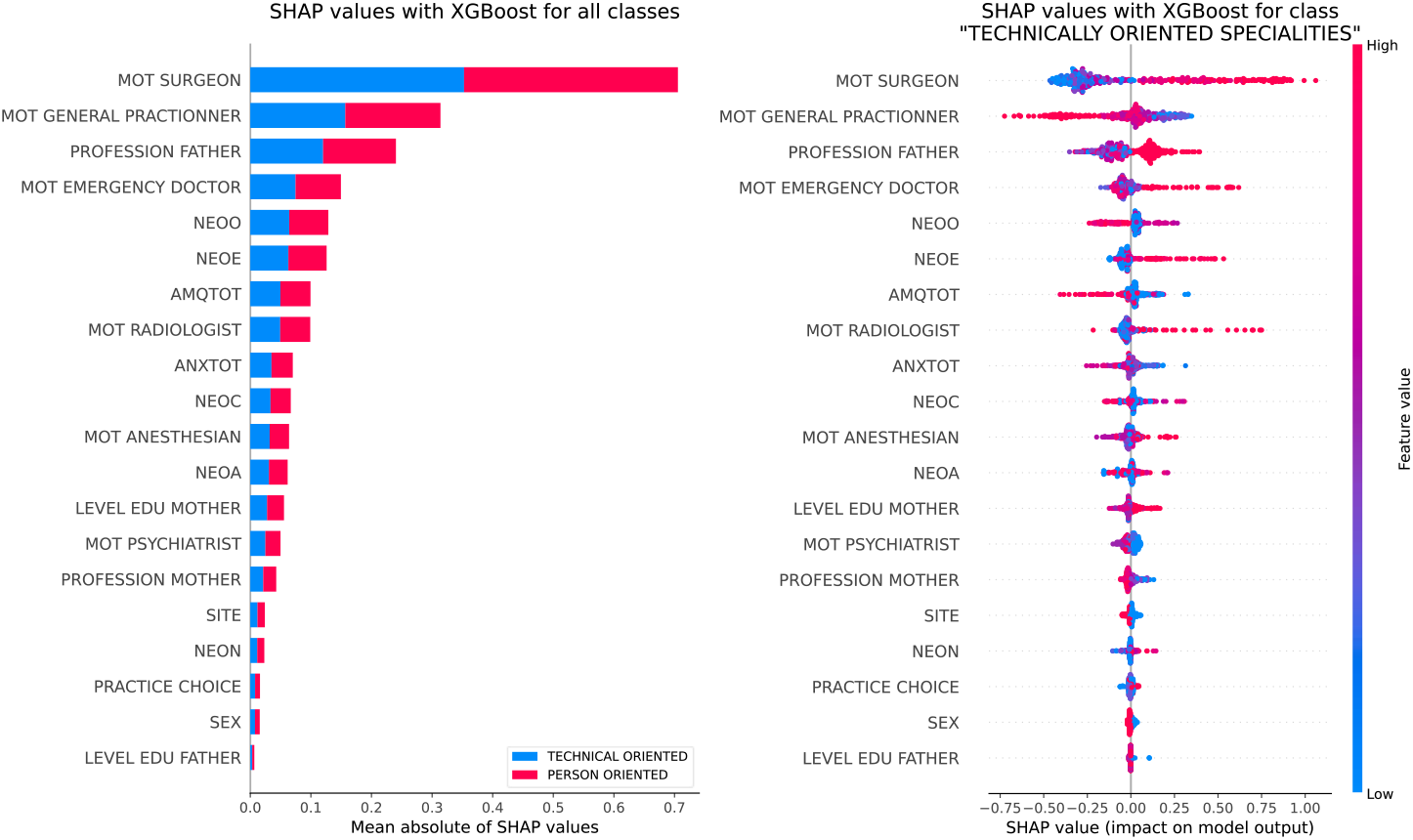
SHAP values with XGBoost classifier.

## 4. Conclusion

In conclusion, this study investigates several machine learning models for predicting medical students’ career choices using data from Swiss and French medical faculties. Gradient-boosting methods achieved the best performance, with accuracy as high as 69%, for predicting between technically and non-technically oriented specialties. Post-hoc interpretability methods highlight the importance of motivation and early specialty preferences in these decisions. Despite these promising results, there is important room for performance improvement, which could be achieved by increasing the training set size and integration of exogenous information. Future work could also focus on investigating the predictive performance of algorithms for more fine-grained career choices.

## Data Availability

Data is not available

